# Genomic dissection of sleep archetypes in a large autism cohort

**DOI:** 10.1101/2025.04.04.25325272

**Authors:** Leo Brueggeman, Natalie Pottschmidt, Tanner Koomar, Taylor Thomas, Jacob J. Michaelson

## Abstract

Poor sleep is a major concern among individuals with autism and their caregivers. To better characterize the genetic and phenotypic heterogeneity of poor sleep in autism, we recruited 5,686 families from SPARK, a nationwide genetic study of autism, who described their sleep experiences using the Children’s Sleep Health Questionnaire (CSHQ) and other self-report items. The collective experiences from this large sample allowed us to discover eight distinct archetypes of sleep in autism. Membership in some of these archetypes showed significant SNP-heritability (0.50 - 0.65, 95% confidence interval = 0.08 - 1), and polygenic estimates of educational attainment, BMI, and ADHD risk contributed extensively to the genetic signatures of these sleep archetypes. Surprisingly, polygenic estimates of general population sleep phenotypes showed sparser and more modest associations, perhaps suggesting that the genetic drivers of disordered sleep in autism may be distinct from those encountered in the general population. GWAS on archetype membership yielded no genome-wide significant loci, however, the most significant gene for the most severe archetype was the nitric oxide (NO) signaling gene *NOS1AP*, which was previously linked to sleep disruption in schizophrenia. Finally, the eight sleep archetypes showed specific signatures of treatment response across five major categories of sleep aid, pointing to the potential of treatment plans that are tailored to the nature of the sleep problem. These findings provide critical new insight into the comorbidities, subtypes, and genetic risk factors associated with disordered sleep in autism.

## 1 Introduction

Autism spectrum disorder (ASD) is a neurodevelopmental condition characterized by restricted, repetitive patterns of interests or behaviors and deficits in social communication, starting from a young age. Although these core features of ASD are sufficient to warrant a diagnosis, people with autism are highly likely to have at least one other co-occurring condition (COC) [1]. These COCs encompass a wide range of frequently comorbid diagnoses like epilepsy, attention-deficit/hyperactivity disorder (ADHD), anxiety, sensory sensitivity, and gastrointestinal upset, among others. One of the most common and distressing comorbidities in autism is problematic sleep behavior. Seen in up to 70% of children [2] with autism, sleep problems are not only associated with a significantly decreased quality of life for the person with autism [3], but have also been found to have a significant impact on their family [4].

The relationship between disordered sleep and autism is highly studied but not well understood. It has been suggested that sleep problems in infancy precede autism development later in childhood [5], but other studies have found that sleep problems arise simultaneously with ASD and then are consistent or worse as the child grows older [6][2]. Despite the “chicken-or-the-egg” questions of directionality, once autism and sleep problems do co-occur, they display a cyclic relationship such that more severe autism symptoms can exacerbate problematic sleep and vice versa [7]. Children with autism who have trouble sleeping tend to have more problematic behaviors in the daytime [8, 9, 10] and are more severely impacted by core symptoms of autism [11, 12].

One of the leading methods used to characterize sleep problems in ASD is the Children’s Sleep Health Questionnaire (CSHQ) [13]. Consisting of over 30 questions (depending on implementation) relating to sleep health, the CSHQ covers a broad range of sleep symptoms including issues related to categories such as parasomnias, sleep anxiety, and daytime tiredness. These categories were suggested by the original authors of the CSHQ as a way of coherently grouping the questions on the CSHQ. Recently, the CSHQ has started to be used to study sleep issues in ASD. As the CSHQ was not designed for the study of sleep issues of individuals with ASD in mind, there have been efforts to find new groupings of the CSHQ questions which are relevant to ASD [14, 15, 16]. As expected, children with ASD have been shown to score higher on the CSHQ and its subscales than typically developing children [17]. Given the diversity of sleep issues experienced by individuals with ASD, there have recently been efforts to cluster individuals with ASD by their sleep issues [18]. By separating into unstable and stable sleep clusters, associations between intellectual function, communication, and additional symptoms were found. However, this study was performed on N=106 low functioning individuals with autism, which limits the specificity of the resulting clusters. A similar approach applied in a much larger sample would be better able to distinguish low-frequency sleep clusters from non-recurrent individual variation.

Beyond comorbid associations, several studies have focused on etiology and have suggested that disruptions of melatonin production and the circadian rhythm may be to blame for sleep problems in ASD. Autism is associated with decreased overall melatonin production and other disruptions to circadian rhythm from early in development [19] - as early as fetal stages [20]. Some research has suggested that dysfunction in the pineal gland, the primary producer of melatonin, may play a role in autism development due to its production of not only melatonin, but also potentially an endogenous N,N-dimethyltriptamine (DMT), which affects neuroplasticity [21]. Sleep function and ASD have each been investigated from a genetic perspective, although the few studies that have probed their shared genetic etiology have yielded mixed results. For instance, despite multiple research groups examining variations of the Circadian Locomotor Output Cycles Kaput (*CLOCK*) gene, which helps to regulate circadian rhythms, it is not clear whether potentially risk-conferring variants seen in autism are actually related to the condition or to the specific sleep problems seen [22, 23]. One study compared the genetic overlap in ASD and sleep disorders and found several shared genes that contributed risk, including those with functions in melatonin synthesis (*ASMT*), circadian entrainment (*CACNA1C*), dopamineric synapse pathways (e.g., *MAOA*, *SCN1A*), and serotonergic synapse pathways (e.g., *HTR2A*, *SLC6A4*) [24]. However, another study using the Simons Simplex Collection (SSC) dataset examined the assocation between 25 circadian genes and actual sleep behaviors in autism and found no significant relationship [25]. Overall, all genetic studies of sleep in ASD so far have limited their focus to classical sleep and circadian pathways.

The present study aims to delineate the nature of disordered sleep in autism, establish its connection to autism’s core symptoms as well as other comorbidities, and answer basic questions about the role of genetic risk factors. While previous genetic studies of sleep in ASD have focused on pathways known to play a role in sleep [22, 23, 24, 25], we use an unbiased genome-wide approach to explore the full range of possible genetic etiologies. Further, we use archetypal analysis [26] to explore associations with particular forms of sleep dysfunction in autism. To accomplish this, we have recontacted 5,686 families who enrolled in SPARK, the largest genetic study of autism to date, and have obtained detailed information on the sleep characteristics of children and adults in these households. Combining this new data with existing phenotypic and genetic data collected by SPARK yields powerful new insights into the nature of disordered sleep in autism.

## 2 Methods

### 2.1 Sleep cohort

Participants were recruited from the nationwide SPARK study [27] via a Research Match recontact study. In total, 5,686 parents (93.3% mothers) responded to survey questionnaires for themselves and one child with autism (Table 1). In addition to the measures assessed as part of this study (see Measures section below), all families also had available demographic, medical, and core autism behavioral data collected through SPARK. This includes a background history survey taken at enrollment, as well as the Social Communication Questionnaire (SCQ) and the Repetitive Behaviors Scale-Revised (RBS-R). Research match participants had their SPARK medical comorbidity and sleep data combined for analyses. A subset of probands also had complete genetic data (whole exome sequencing and genome-wide array genotyping) available from SPARK for analysis.

**Table 1:** Sleep cohort demographics (probands & parents)

| Name | Probands |  |
| --- | --- | --- |
| | Percent or mean | N or ± SD |
| Age | 11.2 | 5.9 |
| Males | 80.7% | 3877 |
| Females | 19.3% | 923 |
| Caucasian | 81.9% | 4337 |
| CSHQ total score | 71.6 | 15.7 |
| ADHD | 36.1% | 1839 |
| SCQ | 22.5 | 6.9 |
| RBS-R | 34.2 | 19.6 |
|  | Parents |  |
| Age (years) | 41.3 | 8.5 |
| Sex - Males | 16.2% | 958 |
| Sex - Females | 83.8% | 4954 |
| PSQI scores | 16.9 | 2.4 |

### 2.2 Measures

#### 2.2.1 Children’s Sleep Health Questionnaire (CSHQ)

This parent-report sleep screening instrument yields both a total score and 8 subscale scores, reflecting key sleep domains that encompass major medical and behavioral sleep disorders in children. A validation study [13] showed adequate internal consistency (*alpha* = 0.68 for a community sample, and = 0.78 in a sample of children seen at a sleep clinic). Test-retest reliability (two-week interval) was acceptable (*r* ranged from 0.62 - 0.79). The CHSQ has been used in a study of quality of life for people with autism[3]. The CSHQ form we used can be seen in Supplementary Table 1. Questions on our CSHQ form are scored from 1 to 5, giving us greater resolution than the typical CSHQ form (scored 0 to 2), but not allowing direct comparison of CSHQ scores.

#### 2.2.2 Pittsburg Sleep Quality Index (PSQI)

The PSQI [28] is a nineteen-item self-report questionnaire which assesses sleep quality and disturbances in adults over the past month. A global PSQI score is calculated based on scores to seven sub-components. Internal consistency and test-retest (M = 28 day interval) reliabilities were good (*alpha* = 0.83, *r* = 0.85). The test was validated by significantly different PSQI scores between a control group and populations of patients with depression or sleep disorders; additionally, a cutoff score of 5 was established to distinguish good (<5) from poor sleepers with a sensitivity of 89.6% and specificity of 86.5%.

### 2.3 SPARK Genotype data quality control, clustering and imputation

Array genotyping data was generated and processed by SPARK (Freeze three, 2019-09-12), see original publication [29] for details. SNPs were mapped to the hg19 reference of the human genome. Mapping was done with liftOver, from hg38 to hg19. All QC steps carried out with PLINK [30] and R [31], based on the QC process described in [32]. R was used for calculating standard deviations from heterozygozity and population outliers.

Samples and SNPs with missing rates above 20% were removed, then samples and SNPs with missing rates above 5% were removed. This happens in two stages so that highly problematic SNPs and individuals do not cause systemic problems at the more stringent threshold. SNPs with MAF < 5% in the cohort were removed, along with SNPs with a HWE p-value < 1e-10. Sample were removed with missing rate > 5% on any one autosome. Samples with heterozygosity rates more than 3 standard deviations from the mean rate were removed. Samples with more than 3 standard deviations from the mean on any of the first 10 MDS components were removed. Also, samples with ethnic backgrounds not captured in 1,000 genomes, or more subtle admixture were removed.

After QC, samples were clustered based on genotype to be assigned to identify population substructure of the sample. The cohort was merged with the samples from the 1,000 genomes phase 3 data. The combined cohort was clustered into 5 groups, representing the 5 distinct super-populations found in 1,000 genomes. Clustering was based on the first 10 components from multi-dimensional scaling of the combined kinship matrix of the cohort and 1,000 Genomes. The top 20 principal components of each of the 5 clusters of the cohort were calculated separately. These components were used in downstream analyses to correct for population substructure. Samples which clustered with the 1,000 Genomes Europeans were used for all subsequent analyses. In total, 194.410 SNPs were removed (out of 616,321 SNPs), along with 1,922 individuals removed due to QC filters.

Samples and SNPs which passed QC were imputed to the 1,000 genomes phase 3 reference using the genipe pipeline [33]. In brief, genipe performs: LD calculation and file handling with PLINK [30], phasing of genotypes done with SHAPEIT [34], and imputation by IMPUTE2 [35].

### 2.4 Heritability and LD-score regression analyses

Estimation of heritability of the CSHQ total score and individual questions was performed using the GCTA GREML software [36]. Heritability analyses were run on SPARK probands of european descent. Individuals with cryptic relatedness > 0.025 (as per GCTA’s documentation) were removed. Heritability analyses were adjusted for an individuals sex, age, age squared, and the top 20 common variant principal components. Heritability results can be seen in Supplementary Table 2.

Linkage-disequilibrium score regression [37] was performed. The heritability and coheritability between ASD [38], 18 sleep phenotypes, and several PGC psychiatric disorders were tested (see Supplementary Table 3). Results from this analysis can be found in Supplementary Table 4.

### 2.5 Common variant association and analysis

The BOLT-LMM [39] software was used to generate genome-wide summary statistics of CSHQ association. As described above, samples were subject to QC and ethnic background filtering, resulting in a total of 2,331 individuals used for genetic association. Associations were controlled for the effect of age, age squared, sex and genetic PCs (n = 20). The resulting summary statistics (Supplementary Table 5) were filtered for MAF < 0.05 and then analyzed used the SNP2GENE module of FUMA [40]. Default parameters were used with the addition of a lead SNP minimum p-value < 0.00001. The full results from the SNP2GENE analysis are available in Supplementary Table 6. A power analysis was performed using the `genpwr` package [41]. The minor allele frequency and effect size thresholds were calculated at 80% power under an additive genetic model (Supplementary Figure 1).

### 2.6 Calculation of polygenic risk scores

Genome wide association study (GWAS) summary statistics were downloaded for common psychiatric conditions and for several sleep phenotypes. The full list and sources of these GWAS studies can be seen in Supplementary Table 3. Polygenic risk scores for all individuals in the SPARK cohort were calculated using the PRSice tool [42]. PRS scores were calculated on imputed genotypes with a minor allele frequency greater than 0.01. Three p-value cutoff values were evaluated, 0.01, 0.1, and 0.5. A linear model was fit to predict total CSHQ score using each PRS p-value cutoff independently. Covariates included in this model correct for the effects of the top 20 genetic principal components, age, and sex. PRS scores were z-scaled prior to modeling. Coefficients and p-values from the 0.01 cutoff were used to create figure 2. These p-values were FDR corrected for the total number of tests (number of PRS by 3 cutoffs). All values for this analysis can be found in Supplementary Table 7.

**Figure 1:**
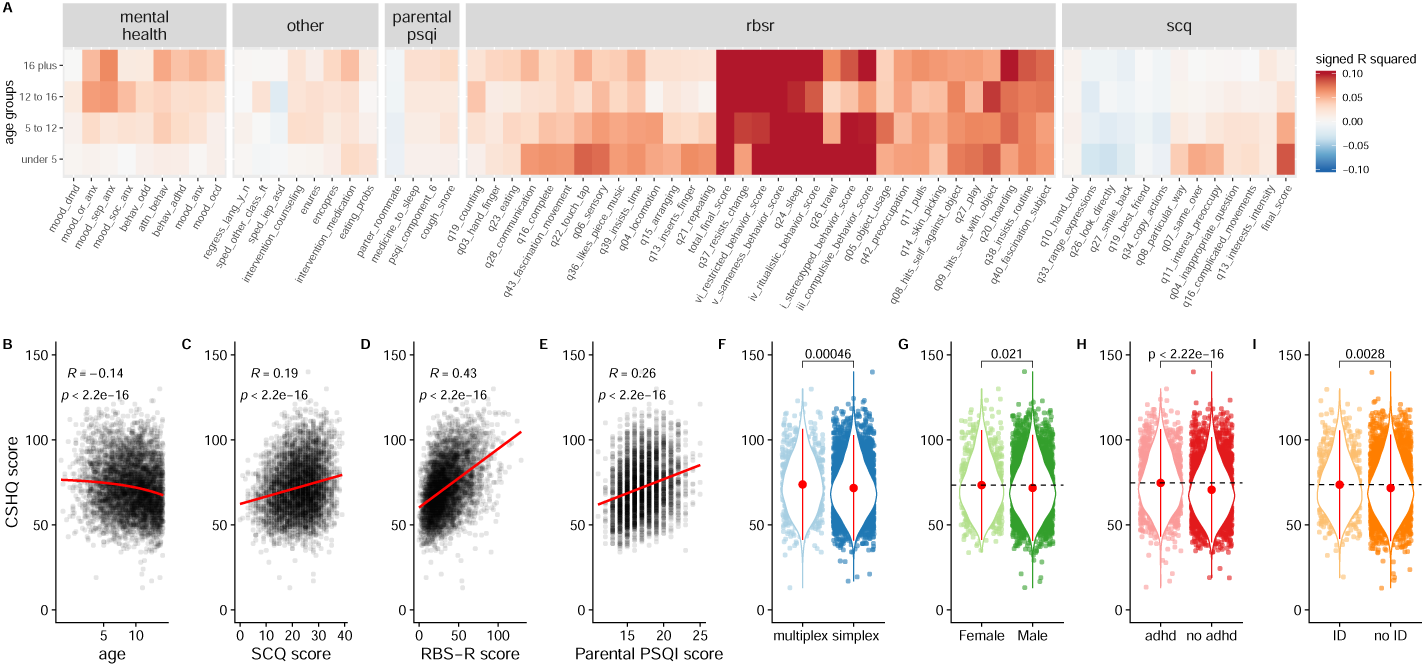
SPARK cohort CSHQ associations. Correlation values (R squared) of comorbidity, diagnostic, and parental sleep questionnaire data with proband total CSHQ score are shown for SPARK participants (n=5686) split by age group (A). The association between total CSHQ score and major diagnostic and demographic factors are shown (B-I).

**Figure 2:**
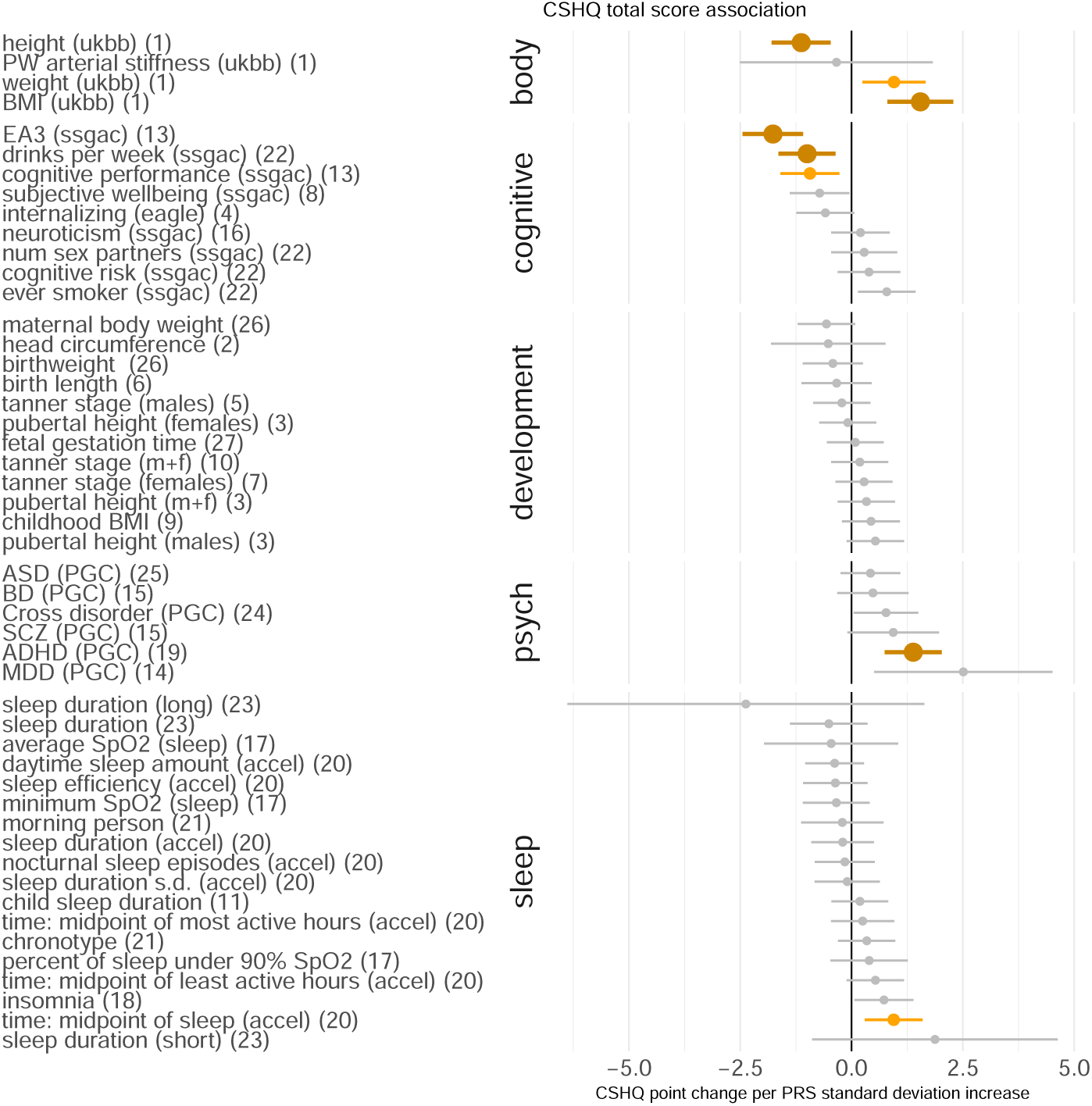
Associations between total CSHQ score and polygenic risk scores (PRS) in ASD. A total of 50 z-scaled PRS scores (SNP p-value threshold = 0.01) were tested for association with total CSHQ score using a linear model. The x-axis represents the coefficients from this model fit. Bolded, dark orange PRS are associated at FDR < 0.05, while lighter orange PRS are associated at FDR < 0.10.

### 2.7 Archetypal sleep analysis

Although the CSHQ includes a number of subscales, they were derived through intuitive domain expertise, and were not data-driven (e.g., from a factor analysis)[13]. Furthermore, our initial analysis showed extreme correlation between these subscales and the CSHQ total score, limiting the specific interpretability of any specific subscale. Finally, the original sample size for the development of the CSHQ (*N* 130) is dwarfed by the sample size of the current study (*N =* 5, 686). Taken together, we felt these points justified a thorough, data-driven assessment of the latent factors underlying sleep in this large sample. Data from the individual CSHQ questions was used for archetypal analysis [43]. CSHQ question responses were residualized for the effect of age and then used as input for archetypal analysis. An optimal *k* was picked by the elbow plot method [44]. To make the archetypes more robust against the stochastic nature of the algorithm, we performed 10 runs of the procedure with *k =* 8 archetypes. The archetypes from each run were then pooled and clustered with the PAM algorithm [45], using *k =* 8 to match the number of archetypes. The medoids of these clusters were then used as the starting point for a final run of archetypal analysis with *k =* 8. The coefficients for the *k =* 8 archetypes are a score for membership in each archetype or cluster. For analyses where a binary assignment to an archetype was desirable (see Fig. 3C), we considered individuals whose maximal archetype coefficient was greater than twice the next-highest coefficient to be examples of “archetypal sleep”, and they were assigned to their maximal-scoring archetype. All other individuals were considered to be mixtures of the archetypes.

**Figure 3:**
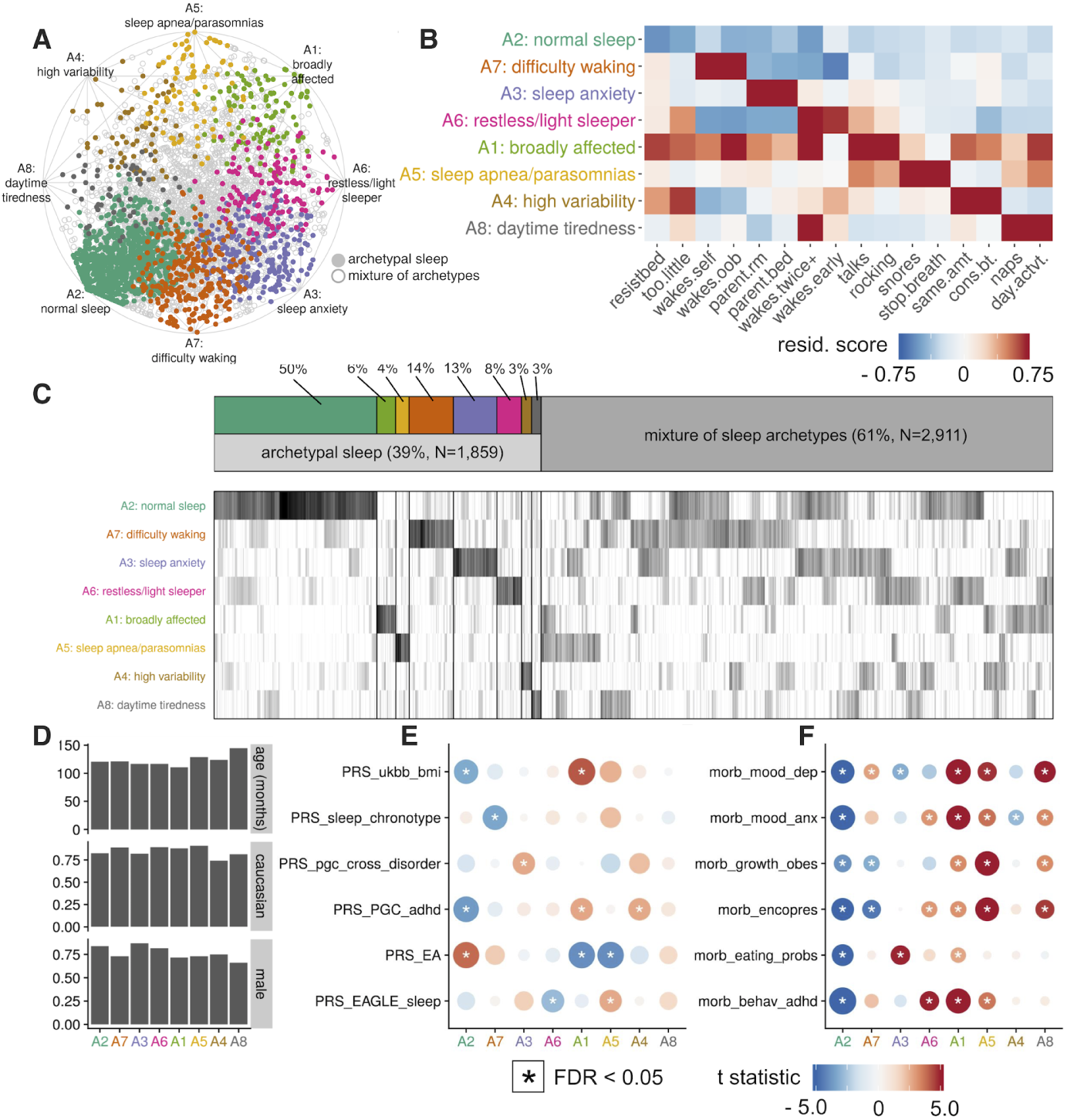
ASD sleep archetypes. Archetypal analysis was performed using CSHQ questionnaire items (A). Characteristic patterns across items of the CSHQ distinguished the sleep archetypes (B). Individuals have a coefficient for each archetype, with some fitting unambiguously (see methods) into a single sleep archetype (C), while most individuals appeared as a mixture of archetypes. Archetypes do not vary drastically in age, sex, or race (D). Sleep archetypes display specific patterns of PRS association (E), with normal sleep (A2) showing depleted risk for high BMI and ADHD coupled with high propensity for educational attainment, and broadly affected (A1) showing the inverse pattern. Sleep archetypes showed specific patterns of comorbidities, most not explicitly related to sleep (F).

### 2.8 Analysis of responsiveness to sleep aids

Participants were asked whether their dependent child took any of five classes of sleep aids (melatonin, antihistamines, PM variants, antidepressants, and non-benzodiazapenes), whether the sleep aid was effective for sleep, and whether any side effects were experienced. We coded a four-level ordinal response as 1: no response and side effects experienced, 2: no response and no side effects experienced, 3: response and side effects experienced, and 4: response and no side effects experienced. We fit a univariate ordinal regression model for each combination of sleep aid (*N =* 5) and sleep archetype (*k =* 8), for a total of 40 models, where archetype coefficients were used to predict response to the sleep aid. *Z* statistics and *p* values were extracted from these model fits to characterize the tendency of response for individuals mapping to the archetypes (Figure 5).

## 3 Results

### 3.1 Phenotypic associations with disordered sleep in ASD

Using the total score from the CSHQ, the association with 320 other phenotypic variables was measured. These variables included measures from a diverse set of sources, including demographics, past medical history, and the SCQ, RBS-R, and PSQI questionaires. Association was measured by independently fit linear regression models of the total CSHQ score (dependent variable) predicted by the 320 phenotypes (independent variable). Data was split into four major age groups of under 5 years old, 5 to 12 years old, 12 to 16 years old, and 16 years old to 17 years old, with 545, 2775, 1060, and 390 observations, respectively. Difference between age groups for the phenotypes was measured by ANOVA or a *χ*-squared test. Associations with sleep score were seen in every class of phenotypic variable tested (Figure 1A), with 70 phenotypes showing significant association with age group and total CSHQ score (Bonferroni threshold: p < 3.4 *×* 10*^−^*^5^).

Among associated variables, age had a highly significant effect on total sleep score (p < 2.2 *×* 10*^−^*^16^, Pearson’s r = −0.14), with each year of life resulting in a 0.57 point decrease on the CSHQ. Autism severity, as measured by the SCQ, also showed a highly significant (p < 2.2 *×* 10*^−^*^16^, Pearson’s r = 0.19) positive correlation with total sleep score. The repetitive behavior scale (RBS-R) was among the variables with highest correlation with total sleep score (p < 2.2 *×* 10*^−^*^16^, Pearson’s r = 0.43). The association with parent-reported sleep scores was also significant (p < 3.7 *×* 10*^−^*^11^, Pearson’s r = 0.26). Lastly, we tested the association with CSHQ of several characteristics thought to broadly impact autism associated phenotypes: family type (simplex or multiplex), reported sex, ADHD, and intellectual disability comorbidity. Significant differences by Mann-Whitney U test were seen in all of these categories, with multiplex families, individuals reporting a female sex status, individuals with ADHD, and individuals with intellectual disability having significantly higher CSHQ scores (p = 0.00046, p = 0.021, p < 2.2 *×* 10*^−^*^16^, p = 0.0028, respectively). The full table of associations can be seen in Supplementary Table 8.

### 3.2 Genetic correlation between ASD and sleep traits

To test for a possible genetic association of sleep traits with ASD, we used LD score regression to compare GWAS summary statistics from studies of ASD[38], sleep (18 sleep traits from 6 individual studies [46, 47, 48, 49, 50, 51]), and several psychiatric disorders. Six sleep traits had a significant genetic correlation with ASD after FDR correction (FDR < 0.05). Specifically, the timing of the midpoint of sleep (*r_g_* = 0.30, SE = 0.07, p=4.01 *×* 10*^−^*^5^, FDR < 0.05) and the m10 timing of sleep (*r_g_* = 0.25, SE = 0.06, p=4.35 *×* 10*^−^*^5^, FDR < 0.05) were both positively correlated with ASD. Conversely, morningness (*r_g_* = −0.18, SE = 0.03, p=4.45 *×* 10*^−^*^7^, FDR < 0.05), chronotype (*r_g_* = −0.19, SE = 0.03, p=8.27 *×* 10*^−^*^8^, FDR < 0.05), sleep duration (*r_g_* = −0.24, SE = 0.05, p=1.36 *×* 10*^−^*^5^, FDR < 0.05), and sleep efficiency (*r_g_* = −0.18, SE = 0.05, p=1.59 *×* 10*^−^*^3^, FDR < 0.05) were all significantly negatively correlated with ASD (Supplemental Figure 2, Supplemental Table 9).

### 3.3 Polygenic predictors of sleep disruption severity in ASD

Given the observed significant genetic correlation between ASD and many sleep traits, we next turned our attention to testing whether polygenic risk for any of these traits would have an impact on sleep disruption in an actual cohort of individuals with ASD. In addition to modeling CSHQ total score using the aforementioned sleep PRS, we also included 49 other psychiatric, cognitive, body morphometry, and cognitive-related traits.

The PRSice software was used to extract polygenic risk scores for each trait at the p-value cutoff points of 0.01, 0.1 and 0.5. A linear model was independently fit to predict the total sleep score (dependent variable) using each genetic predictor (independent variable), at each cutoff, and the covariates (20 genetic PCs, age, age squared, sex). The coefficients from this model fit using the 0.01 cutoff point are shown in Figure 2. Polygenic risk scores were z-scaled, so a 1 unit increase in PRS risk can be interpreted as an increase by one standard deviation of the PRS score. A total of 5 polygenic predictors survived FDR correction, after accounting for the number of traits and cutoff points tested. These included BMI (*B* = 1.54, SE = 0.37, p = 4.3 *×* 10*^−^*^5^, FDR < 0.05), height (*B* = −0.94, SE = 0.34, p = 5.9 *×* 10^3^, FDR < 0.05), educational attainment (*B* = −1.76, SE = 0.34, p = 3.8 *×* 10*^−^*^7^, FDR < 0.05), ADHD (*B* = 1.38, SE = 0.32, p = 2.4 *×* 10*^−^*^5^, FDR < 0.05), and drinks per week (*B* = −0.99, SE = 0.32, p = 2.2 *×* 10*^−^*^3^, FDR < 0.05). Additionally, 3 polygenic predictors were nominally significant, including weight (*B* = 0.95, SE = 0.36, p = 8.3 *×* 10*^−^*^3^, FDR < 0.1), cognitive performance (*B* = −0.93, SE = 0.33, p = 5.7 *×* 10*^−^*^3^, FDR < 0.05), and the midpoint of sleep (*B* = 0.94, SE = 0.33, p = 4.3 *×* 10*^−^*^3^, FDR < 0.05).

### 3.4 Sleep archetypes in ASD with unique genetic and comorbidity associations

To test for the possibility of distinct subtypes of disordered sleep in ASD, we used archetypal analysis [43]. In total, 8 archetypes were identified which lie along the convex hull of the multi-dimensional CSHQ data (Figure 3A). These archetypes have unique profiles in the age-residualized CSHQ questions (Figure 3B). Archetypes 1 and 2 correspond to broadly affected and normal sleep, respectively, while archetypes 3, 4, 5, 6, 7 and 8 correspond to more specific problems being enriched within the CSHQ. Assigning individuals to these archetypes (see methods) showed that 39% of the sample was specifically enriched for one of the archetypal CSHQ patterns, with the remaining being a combination of several archetypes (Figure 3C).

Using archetype assignment scores, we then tested associations with polygenic risk scores and comorbidities. Analysis of the individuals uniquely assigned to each archetype (N = 1,859) showed similar profiles in age, sex, and race (Figure 3D). Using archetype assignment scores across the full sample showed a number of significant PRS associations (Figure 3E). Educational attainment (*B* = −0.011, t = −3.69, p = 2.3 *×* 10*^−^*^4^, FDR = 1.7 *×* 10*^−^*^3^) and ADHD (*B* = −0.020, t = −3.53, p = 4.1 *×* 10*^−^*^4^, FDR = 2.8 *×* 10*^−^*^3^) had the most significant PRS associations with archetype 5 and 2, respectively. There were also many significant associations with comorbidities (Figure 3F), with encopresis, anxiety, and depression having associations with six of the eight archetypes (FDR < 0.05). The full list of archetype associations can be found in Supplementary Table 10.

### 3.5 Heritability of sleep disruption in ASD

To further investigate the potential genetic basis of sleep disruption in ASD, as measured by the CSHQ, we used the GCTA GREML [36] tool to study the SNP-heritability of total sleep score in individuals with ASD with shared genetic backgrounds (N = 1957). SNP-heritability (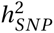) of the total sleep score was estimated to be 0.65, with a standard error of 0.21 (p = 0.0012, FDR = 0.01). Additionally, three sleep archetypes (Figure 4A) showed at minimum nominal significance (FDR < 0.1), including: archetype 1 (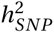 = 0.53, SE = 0.21, p = 0.0082, FDR = 0.05), archetype 2 (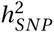 = 0.65, SE = 0.21, p = 0.0011, FDR = 0.01), and archetype 3 (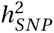 = 0.50, SE = 0.21, p = 0.0085, FDR = 0.05).

**Figure 4:**
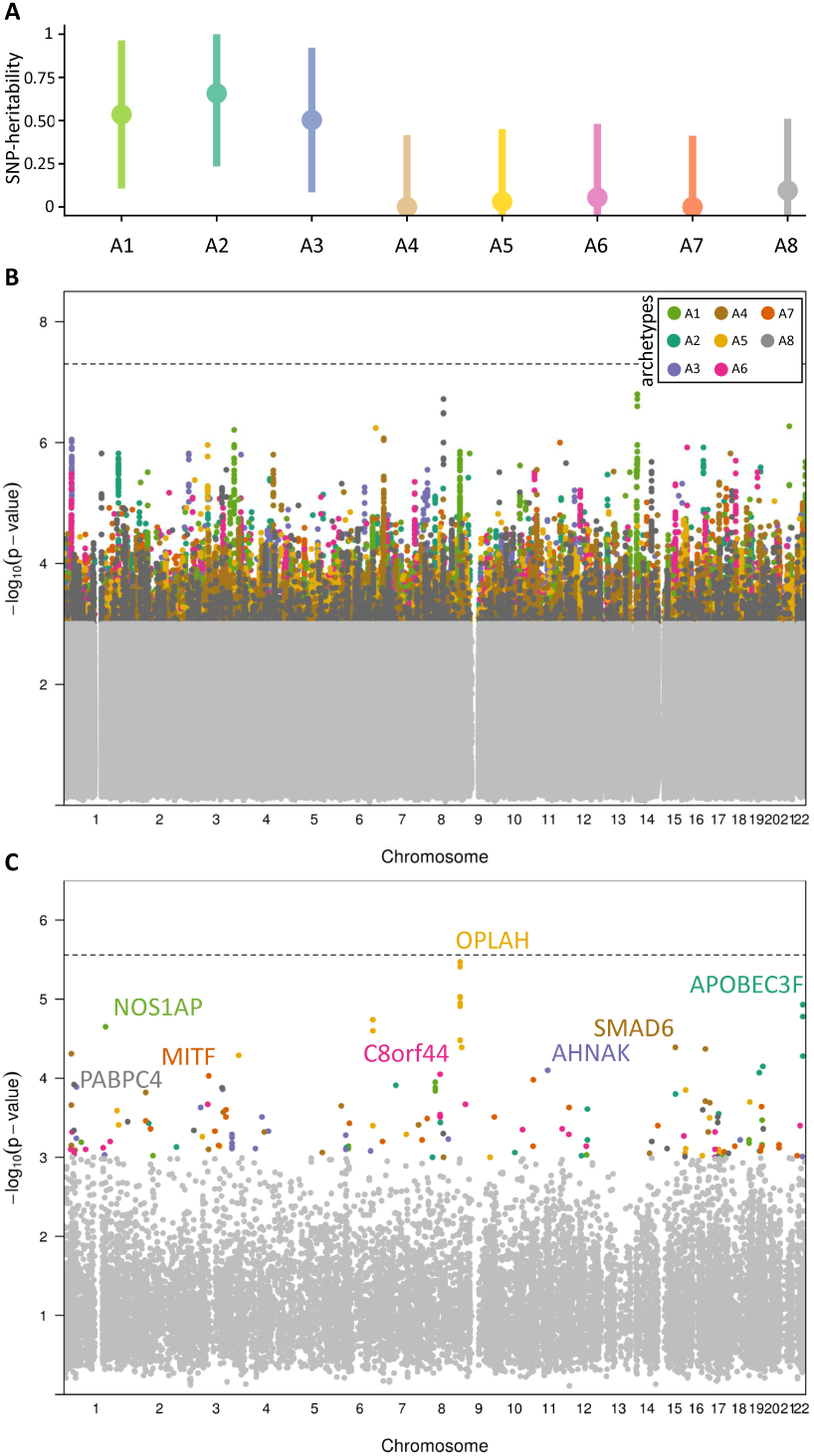
Heritability, GWAS, and MAGMA analysis of sleep archetypes. GCTA-GREML heritability estimates of the sleep archetypes (A). GWAS on sleep archetype coefficients with SNP color representing the archetype with the most significant p-value (B). MAGMA gene-level analysis of archetype GWAS results, with the same coloring scheme as the SNP plot (C). The top gene for each archetype is shown. Points below significance level of p=0.001 are shaded light grey.

Beyond total score and sleep archetypes, individual CSHQ questions were also tested for SNP-heritability. Across 32 ordinal questions on the CSHQ, 2 questions were found to be significantly heritable after FDR correction. Of these questions, waking up in a negative mood had the most significant SNP-heritability of 0.69, with a standard error of 0.21 (p = 0.0006, FDR = 0.01). The other question having significant heritability was sleeping in parents bed (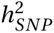 = 0.65, SE = 0.21, p = 0.0009, FDR = 0.01).

### 3.6 Common variant associations with ASD sleep archetypes

A genome-wide association study was performed for the total CSHQ score and sleep archetypes and analyzed using the FUMA software. A table of the top SNPs from each of these phenotypes is available (Supplementary Table 5). None of the SNPs (Figure 4B) reached genome-wide significance (*p <* 5 *×* 10*^−^*^8^).

We used the SNP2GENE module in FUMA[40] to summarize archetype GWAS results at the gene level (Figure 4A). No genes reach genome-wide significance after accounting for multiple testing (Figure 4C). *NOS1AP* was among the top genes identified by MAGMA (Figure 4C) [40], and was associated with the most severe sleep archetype (archetype 1).

### 3.7 Sleep aid responsiveness in ASD

To compare sleep aids used in ASD, we collected parent response questionnaire data on the effectiveness of several major sleep aid classes. Out of all drug classes assessed, melatonin was most widely used, with 59% of the sample having taken melatonin at least once (Figure 5A). To assess the effectiveness of different sleep aids between the sleep archetype groups (N = 1,859) we fit ordinal regression models (one for each combination of sleep aid and archetype) predicting overall responsiveness to the sleep aid. Melatonin’s response profile was significant across seven of the eight sleep archetypes, with some archetypes showing resistance to melatonin (Figure 5B), and some archetypes increasing the probability of response to melatonin (Figure 5C). Archetype 1 (broadly affected) showed the most treatment resistance among all archetypes (Figure 5D).

**Figure 5:**
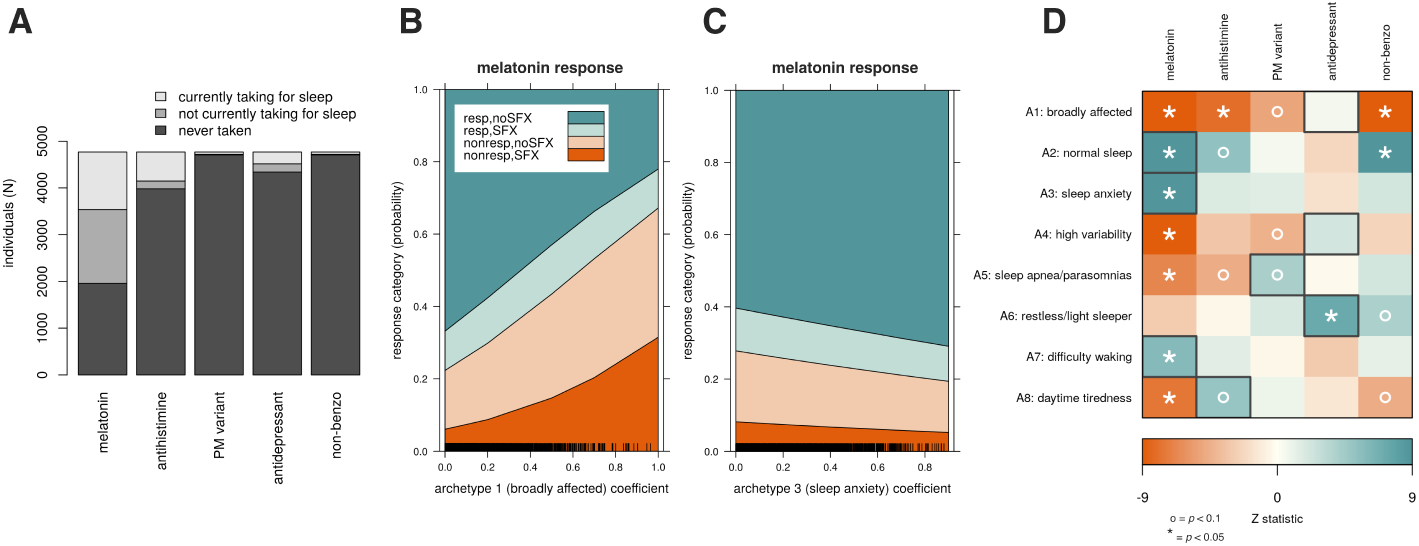
Response to sleep aid varies by sleep archetype. Number of individuals who have taken each category of sleep aid (A). Some archetype coefficients are associated with decreasing sleep aid response, e.g., melatonin (B), while others are associated with increasing response (C). We performed this analysis over all combinations of archetype and sleep aid (D). Increased responsiveness is indicated with a blue-green shade, while decreased responsiveness is indicated in orange.

## 4 Discussion

Poor sleep profoundly impacts quality of life for many individuals with autism, and our limited understanding of the relationship between sleep and autism itself stands in the way of effective and individualized interventions. By recontacting 5,686 families who participated in the SPARK study [27], we used an already well-characterized cohort to map the landscape of disordered sleep in autism. While sleep disturbance in ASD has been extensively studied [52], including an impressive, large-scale phenotypic study focusing only on the CSHQ [14], no large-scale, genome-wide study on sleep in ASD has been performed. Previous genetic studies of sleep disturbance in ASD limited their search space to genes known to have a role in the circadian rhythm pathway [53, 23, 54, 25]. In this study, we expanded to a genome-wide search space and found compelling evidence for genetic factors in the diverse presentation of sleep disruption in ASD.

We found that overall sleep disruption, as measured by the total score from the Children’s Sleep Health Questionnaire (CSHQ), increases significantly with higher SCQ (Pearson’s r = 0.19, p < 2.2 *×* 10*^−^*^16^) and RBS-R (Pearson’s r = 0.43, p < 2.2 *×* 10*^−^*^16^) scores (see Figure 1). Although this would seem to suggest that the core symptoms of autism are intrinsically connected to disordered sleep, we instead found that polygenic risk for ADHD, and not for ASD, was significantly associated with disrupted sleep (Figure 2). This association was robust even after inclusion of diagnosis with ADHD as a covariate (*p =* 0.001, *t =* 3.29). Other elements of polygenic risk, including height, BMI, educational attainment, and drinks per week showed significant phenotypic correlations in agreement with previous literature [55, 46]. Interestingly, although these polygenic risk factors predicted poor sleep as they had in other genetic studies of sleep, the polygenic risk estimates for the sleep traits themselves, largely derived from disorder-agnostic studies, were not predictive of disrupted sleep in our sample (Figure 2; note midpoint of sleep as the lone exception). This may suggest that the genetic risk factors that underlie sleep disruption in autism might either be conditional on the presence of disrupted neurodevelopment, or that the kinds of disordered sleep experienced in autism are simply not sufficiently represented in these general population studies. Alternatively, insufficient power in these previous studies, or in the current study, may contribute to the observed lack of association (although we found evidence of significant heritability for the CSHQ total sleep score in our sample). In any case, we find little evidence that the knowledge gained from previous studies on the genetics of sleep is applicable to disrupted sleep (broadly measured) in autism.

Given that this is the largest genetically informative sample of sleep-phenotyped individuals with autism, we took the opportunity to search for evidence of specific recurrent patterns of disrupted sleep. Using archetypal analysis, we found support for *k =* 8 distinct sleep archetypes in autism (see Figure 5), which, from a conceptual perspective, are congruent with the subscale definitions that were originally (though somewhat arbitrarily) defined for the CSHQ [13]. About 40% of our sample mapped unambiguously to one of these sleep archetypes, while 61% of the sample presented as a mixture of two or more archetypes. About 20% of our overall sample mapped unambiguously to archetype 2 (normal sleep), suggesting that as many as 80% of individuals with autism show signs of disordered sleep. The most severe sleep disruption was seen in archetype 1, which we called “broadly affected” because of its pervasive endorsement of a variety of sleep problems (see Fig. 3B). About 6% of individuals with archetypal sleep patterns were mapped to this archetype, or about 2.4% of the total sample. These individuals tend to resist bedtime, need appreciable parental input to go to bed (e.g., rocking), wake during the night, talk during sleep, have difficulty getting out of bed, and then sleep during normal daytime activities. When considering polygenic risk (PRS) associated with this archetype, we found significant negative association with educational attainment and positive associations with ADHD and BMI (all *FDR <* 0.05, see Figure 3E). Parental endorsement of the child’s obesity and ADHD diagnosis were congruent with these PRS associations (Fig. 3F), and additionally showed strong concern about depression and anxiety compared to other sleep archetypes.

While archetypes 1 (broadly affected) and 2 (normal sleep) cover the extremes of the sleep patterns seen in our sample, six other archetypes captured more distinct forms of sleep disruption in ASD. Archetype 3 (sleep anxiety, 13% of archetypal sleep, 5.2% of total) was characterized by a need for parental proximity and reassurance as part of routine sleep. This archetype distinguished itself from other sleep archetypes through its significantly increased PRS for the PGC cross-disorder trait, suggesting excess neuropsychiatric burden compared to other archetypes, as well as increased parental endorsement of eating problems. Archetype 4 (high variability, 3% of archetypal sleep, 1.2% of total) was characterized by constantly shifting bedtimes and sleep duration, with a parental impression of too little sleep. Aside from archetype 1, this was the only other archetype to show a significant positive association with ADHD PRS. Archetype 5 (sleep apnea/parasomnias, 4% of archetypal sleep, 1.6% of total) showed pronounced presence of breathing issues (snoring, apnea) as well as sleep talking. Parent report showed strong endorsement of encopresis and obesity, as well as ADHD, depression, and anxiety. Archetype 6 (restless/light sleeper, 8% of archetypal sleep, 3.2% of total) was defined by waking repeatedly in the night and waking too early. Archetype 7 (difficulty waking, 14% of archetypal sleep, 5.6% of total) was characterized by parent report of having to wake the child, coupled with difficulty getting out of bed. This was the only archetype to show a significant PRS association with chronotype (negative association or “night owl” chronotype). Finally, archetype 8 (daytime tiredness, 3% of archetypal sleep, 1.2% of total) was characterized by waking repeatedly at night and then needing to take naps during the day, as well as sleeping during normal daytime activities. This was the only archetype that did not show any significant PRS associations for the traits we examined, although there was significant enrichment of anxiety, depression, obesity, and encopresis as comorbidities. Together, these results underscore the heterogeneity in both presentation and etiology of the diverse forms of sleep disruption encountered in ASD while providing a map for navigating it.

Although our sample was not well-powered for GWAS and no associations survived multiple testing correction, gene-wise MAGMA results (see 5C) pointed to potentially interesting genes, including *NOS1AP*, which was the top gene for archetype 1, the most severely affected form of sleep disruption we observed. *NOS1AP* is involved in nitric oxide signaling and was previously implicated in disrupted sleep in schizophrenia[56], as well as autism itself [57]. Nitric oxide (NO) signaling, facilitated by nitric oxide synthases, is critical for sleep, and experimental depletion of NO in animal models completely abolishes the capacity for sleep [58].

Our archetypal analysis allowed us to interpret response to several major classes of sleep aid with greater granularity (i.e., compared to the CSHQ total sleep score), and we were able to tell, for each sleep archetype, which sleep aids were likely to work well, and which showed little evidence of efficacy in that context (see Figure 5). As has been previously reported in autism[59], the most widespread sleep aid used among our sample was melatonin, followed by antihistamines (Fig. 5A). Interestingly, we found that all sleep archetypes had at least one significant association (*p <* 0.05) with sleep aid response, some tending toward lower responsiveness (Fig. 5B) and some to higher (Fig. 5C). This indicates that these sleep archetypes may be able to steer sleep aid choice, when better clinical predictors are not available. Given the limited number of sleep aid trials in ASD [60], this may be an important first step towards creating more individualized sleep aid treatment plans, especially for those sleep archetypes where no sleep aid shows effectiveness (Fig. 5D).

Our study’s large sample size and availability of genetic data have yielded important new insights into the nature of disordered sleep in autism. We showed the structure and heterogeneity of sleep problems in autism through archetypal analysis. We found that genetic studies of sleep issues in the general population translated only weakly to our autistic sample. We also showed a general lack of evidence for a central role of known circadian genes in disordered sleep in ASD. Instead, we found strong polygenic associations with BMI and ADHD for some archetypes, and neuropsychiatric risk for others. We also found that patterns of sleep aid responsiveness vary extensively according to sleep archetype and may eventually inform individualized interventions.

Despite these advances, the subjective parent-report nature of the data and the dynamic qualities of sleep problems are inherent limitations that make it difficult to understand the nature of the disrupted sleep with much precision. For instance, although we instructed respondents to report their current experiences, several parents related to us that if we had asked them last year, they would have responded to certain items differently. Furthermore, although our sample size breaks new ground for studies of this kind, we will need additional participants to transition this work from the realm of confirming a role for genetics to pinpointing specific genes and potential mechanisms. Our power analysis suggests that we will begin to see productive GWAS results at N > 10,000.

Equipped with these new insights about the existence and nature of the sleep archetypes we present here, future work should focus on more specific investigations, e.g., of one or more of the sleep archetypes, their effective management, and long-term outcomes.

## Data Availability

All data produced in the present study are available upon reasonable request to the authors.

## Acknowledgments

We are grateful to all of the families in SPARK, the SPARK clinical sites and SPARK staff. We appreciate obtaining access to genetic and phenotypic data for SPARK data on SFARI Base. Approved researchers can obtain the datasets described in this study by applying at https://base.sfari.org. This work was supported by the National Institutes of Health [MH105527 and DC014489 to JJM]. This work was supported by a grant from the Simons Foundation (SFARI 516716, JJM). This work was funded in part by the Department of Psychiatry at the University of Iowa, the University of Iowa T32 Genetics Predoctoral Training Program to LB, and the NASA Iowa Space Grant Consortium to TK.

